# Durability and Seasonal Variation in the Effectiveness of Nirsevimab over Three Seasons in Connecticut

**DOI:** 10.64898/2026.06.22.26356264

**Authors:** Hanmeng Xu, Camilla Aparicio-Llorente, Joshua L. Warren, Lee Kennedy-Shaffer, Virginia E. Pitzer, Daniel M. Weinberger, Carlos R. Oliveira, the PROVE-ID Group Authors

## Abstract

**Background:** Nirsevimab has been widely administered in the United States since 2023 to protect infants and young children from severe disease caused by respiratory syncytial virus (RSV). Although early post-licensure studies have shown high effectiveness against medically attended RSV infection, uncertainty remains about the durability of protection, effectiveness beyond the first RSV season, and the extent to which changing RSV seasonality influences real-world effectiveness.

**Objective:** To estimate the effectiveness of nirsevimab against medically attended RSV infection across three consecutive RSV seasons and to examine how effectiveness varies by season and time since immunization.

**Methods:** We conducted a test-negative case-control study utilizing electronic health records of infants and young children tested for RSV by polymerase chain reaction in outpatient and inpatient settings within the Yale New Haven Health System between October 1, 2023, and March 1, 2026. Effectiveness of nirsevimab was estimated using multivariable logistic regression, adjusting for age, weekly RSV activity, pre-existing risk factors, and other potential confounders. Variation in effectiveness was examined by season, encounter setting, and time since immunization up to 24 months.

**Results:** Overall, 17,755 infants and young children were tested for RSV infection, of whom 2,388 (13.4%) were cases and 15,367 (86.6%) were controls. The overall effectiveness of nirsevimab was 67.3% (95% confidence interval [CI]: 59.8, 73.3%) against all medically-attended RSV infections, 60.2% (95% CI: 49.6, 68.5%) against RSV-associated outpatient visits, and 88.9% (95% CI: 82.3, 93.0%) against RSV-associated hospitalization. Effectiveness against medically attended RSV infection declined across seasons, from 76.7% (95% CI: 60.5, 86.3%) in 2023/24 to 54.4% (95% CI: 33.0, 68.9%) in 2025/26. Lower season-specific effectiveness in later seasons corresponded with progressively delayed RSV activity over. Protection against RSV-associated hospitalization declined with increasing time since immunization, from 92.5% (95% credible interval [CrI]: 85.9, 96.4%) at 1 month, to 77.2% (95% CrI: 60.4, 87.6%) at 6 months, and 39.9% (95% CrI: 2.4, 63.3%) at 12 months post-immunization, after which effectiveness plateaued.

**Conclusions:** Nirsevimab remained effective against RSV-associated hospitalization through 6 to 12 months after immunization. Delayed RSV activity was associated with lower effectiveness, highlighting the importance of aligning administration with local RSV circulation.

## Introduction

Respiratory syncytial virus (RSV) is a major cause of lower respiratory tract infection (LRTI) and hospitalization in infants under 1 year old, both globally and in the United States (US) [1,2]. The first extended half-life monoclonal antibody for preventing RSV infections, nirsevimab, was approved in 2023 for routine use in infants and young children. Together with maternal RSV vaccination and newer monoclonal antibody options [3,4], nirsevimab has shown high effectiveness in post-licensure studies [5–9], but knowledge gaps remain about its real-world effectiveness.

A key remaining question is the durability of protection after nirsevimab administration. Post-licensure studies suggest 50%-70% protection against RSV-associated hospitalization up to 7 months after immunization [10,11], but effectiveness beyond 7 months remains unclear. Furthermore, most effectiveness studies have focused on a single RSV season; it is therefore unclear how the effectiveness of nirsevimab varied across seasons since its approval. Timing of RSV activity can vary from year to year; however, the recommendation for seasonal administration of nirsevimab is fixed, with a single dose of antibody administered to infants less than 8 months of age shortly before the RSV season begins (in October-November) and to newborns during the RSV season (October to March). Therefore, the interval between nirsevimab immunization and RSV exposure can also vary, which could lead to lower effectiveness in seasons with delayed RSV activity as protection from nirsevimab wanes over time.

To address these gaps, the primary aim of this study was to evaluate whether season-to-season variation in nirsevimab’s effectiveness was associated with shifts in the timing of RSV activity. Our secondary aim was to characterize the durability of protection by time since immunization.

## Methods

### Data Sources and Study Population

A test-negative case-control design was used to estimate the effectiveness of nirsevimab against medically attended RSV infection, RSV-associated outpatient visits, and RSV-associated hospitalization. The study population included patients aged 36 months and younger who were tested for RSV while receiving care for suspected acute respiratory infection (ARI) in facilities affiliated with the Yale New Haven Health System (YNHHS) between October 1, 2023, and March 1, 2026. The YNHHS is the largest health system in Connecticut, and consists of five integrated hospital networks, 30 emergency or urgent care centers, and more than 130 outpatient clinics in Connecticut; Westchester County, New York; and Rhode Island, all integrated using a single electronic health record (EHR) system. The age cutoff allowed capture of children immunized during infancy who were tested for RSV in subsequent seasons, enabling assessment of effectiveness up to 24 months after immunization. All eligible records were included and we did not exclude patients with multiple RSV tests within or across seasons.

We extracted baseline characteristics, clinical data, RSV test results, and encounter setting from the YNHHS EHR. Baseline characteristics included age, sex, self-reported race and ethnicity, gestational age, birth weight, and insurance type. Respiratory symptoms documented within 14 days of the RSV test and risk factors for severe RSV disease documented in the medical history were identified using free-text searches and ICD-10-CM codes (Table S1). Immunization history was ascertained from CT WiZ, Connecticut’s statewide immunization information system. Electronic reporting to CT WiZ is mandatory for immunizations administered in Connecticut, and CT WiZ includes records available through established electronic data exchanges with immunization information systems in New York and Rhode Island. Maternal RSV vaccination was ascertained by linking infant records to maternal delivery and immunization records when available.

### Study Definitions

Cases were defined as patients with medically attended RSV infection confirmed by nasopharyngeal polymerase chain reaction, and controls were defined as patients with ARI who tested negative for RSV. Analyses were stratified by encounter setting. RSV-associated hospitalization analyses were restricted to patients hospitalized within 14 days of the RSV test. RSV-associated outpatient visit analyses included patients with ambulatory clinic, urgent care, or emergency department encounters that did not result in hospitalization. Patients were classified as immunized if they had documented receipt before the RSV test date. Clesrovimab became available during the study period but was not included in the exposure definition because no participants received clesrovimab.

### Statistical Analysis

Characteristics of the study population were summarized using frequency distributions and measures of central tendency, and compared by RSV case status, season of RSV test, and receipt of nirsevimab. Covariate balance between compared groups was assessed to detect potential confounders using standardized mean differences (SMD), with absolute SMDs of less than 0.20 indicating adequate balance [12–15].

### Effectiveness of Nirsevimab by Season

An overview of the analyses and of the eligibility of the population is presented in Figure S1. We defined the surveillance year as the time period from July 1 to June 30 of the following year and RSV season as October 1 to March 31 of the following year. In our primary analysis, we stratified the study population by surveillance year during which they received the RSV test and estimated the overall effectiveness of nirsevimab for each year. The analytic population for each surveillance year included patients tested for RSV during that surveillance year (July-June) who were also eligible for nirsevimab during that RSV season (i.e., born between October 1 and March 31 of the surveillance year or aged 0 to 8 months at the start of October). Patients who received nirsevimab during the previous RSV seasons were excluded from the primary analysis. Effectiveness of nirsevimab was estimated using multivariable logistic regression models, adjusting for *a priori*-selected confounders including age of patients at RSV test (in months, continuous), RSV activity in the study site (continuous), presence of at least one risk factor for severe RSV disease (binary, definition in Table S1), and maternal RSV vaccine status (categorical: unimmunized, immunized within 6 months of test, immunized over 6 months before test, and unknown). RSV activity was included as a log-transformed continuous variable, derived from the weekly positivity rate of RSV in Connecticut, obtained from the National Syndromic Surveillance Program (NSSP) via the PopHIVE data platform [16]. Effectiveness was calculated as (1 - adjusted odds ratio) * 100%. We conducted six sensitivity analyses in which we: 1) excluded all patients who were eligible for nirsevimab in the previous season; 2) excluded all patients who were coinfected with influenza, SARS-CoV-2, and/or human metapneumoviruses (hMPV); 3) restricted test-negative controls to only those who tested positive for other respiratory viruses; 4) excluded all patients whose mother received RSV vaccine; 5) adjusted for calendar month of RSV test instead of weekly RSV activity; 6) imputed the vaccine status of mothers with unknown vaccine status using the proportions observed among mothers with known status.

To assess whether the initial effectiveness of nirsevimab varied by surveillance year, we estimated the effectiveness among those who were immunized within 2 months before the test, adding an interaction term between immunization and year. The significance of between-year differences in the initial effectiveness was evaluated using a Wald test on the interaction terms.

### Waning Effectiveness of Nirsevimab

In our secondary analyses, the extent to which the protection from nirsevimab varied over time since immunization was estimated using logistic regression within a Bayesian framework, adjusting for the same set of confounders as in the primary analysis [10]. In the waning model, the effect of nirsevimab was allowed to vary by time from immunization to RSV testing. The coefficients for time since immunization were modeled using a spline structure, in which the effectiveness coefficient at each time point was expressed as a linear combination of basis functions capturing smooth underlying trends. We tested several alternative model structures as sensitivity analyses, and a detailed description of all the model structures is provided in the Supplementary Methods. We examined effectiveness over time both by surveillance year and by pooling all three years. Posterior medians and 95% credible intervals (CrI) were used to summarize findings, and model convergence was evaluated using trace plots (Figure S6).

To test whether the shape and initial value of the waning curve varied by surveillance year, we pooled all the records and jointly estimated the difference in effectiveness coefficients between years for each time since immunization interval in one model. Posterior distributions of the differences were plotted and compared to 0 (no difference between years).

All data cleaning and analyses were conducted in R, version 4.3.1[17], with code documented in https://github.com/Hanmeng-Xu/season2_RSV_risk.

## Results

### Study Population

Between October 1, 2023, and March 1, 2026, a total of 17,755 RSV tests were performed among patients aged <36 months in the YNHHS and were included as the study population (Table 1, Figure S1, S2). A total of 3,969 tests were performed in 2023/24 (July-June), 8,249 in 2024/25, and 5,537 in 2025/26. The study population consisted of 2,388 cases (13.4%) with RSV-positive results and 15,367 controls (86.6%) with RSV-negative results. Most RSV tests were conducted in the outpatient setting (n=14,460, 81.4%), and over half of the positive tests occurred during December to January. Age of children at test, gestational age, birth weight, type of insurance, and presence of risk factors were comparable between cases and controls (Table 1). A total of 4,266 patients (24.0%) had ever received nirsevimab in the past, and the immunized patients were more likely to be premature (24.4% vs. 14.7%, SMD = 0.25), to have had lower birth weight (3129 g vs 3249 g, SMD = -0.22), and to have at least one risk factor for severe RSV infection (Table S2).

**Table 1.** Characteristics of cases and controls.

| Characteristic | Overall, N = 17,755<sup>1</sup> | Cases, N = 2,388<sup>1</sup> | Controls, N = 15,367<sup>1</sup> | SMD<sup>2</sup> |
| --- | --- | --- | --- | --- |
| **Sex** |  |  |  | 0.02 |
| Female | 7,685 (43.3%) | 1,052 (44.1%) | 6,633 (43.2%) |  |
| Male | 10,067 (56.7%) | 1,336 (55.9%) | 8,731 (56.8%) |  |
| Missing | 3 (0.02%) | 0 | 3 (0.02%) |  |
| **Race and ethnicity** |  |  |  | 0.12 |
| Black non-Hispanic | 3,303 (18.6%) | 427 (17.9%) | 2,876 (18.7%) |  |
| White non-Hispanic | 4,360 (24.6%) | 669 (28.0%) | 3,691 (24.0%) |  |
| Other non-Hispanic<sup>3</sup> | 1,067 (6.0%) | 133 (5.6%) | 934 (6.1%) |  |
| Hispanic | 8,043 (45.3%) | 1,000 (41.9%) | 7,043 (45.8%) |  |
| Unknown | 982 (5.5%) | 159 (6.7%) | 823 (5.4%) |  |
| **Age at RSV test (months)** |  |  |  | 0.06 |
| Median (IQR) | 10.1 (5.3, 16.5) | 10.2 (5.5, 17.0) | 10.0 (5.3, 16.4) |  |
| **Time of test** |  |  |  | 0.66 |
| 2023/24 RSV season | 3,391 (19.1%) | 661 (27.7%) | 2,730 (17.8%) |  |
| 2024 off-season | 1,713 (9.6%) | 38 (1.6%) | 1,675 (10.9%) |  |
| 2024/25 RSV season | 6,212 (35.0%) | 994 (41.6%) | 5,218 (34.0%) |  |
| 2025 off-season | 2,032 (11.4%) | 30 (1.3%) | 2,002 (13.0%) |  |
| 2025/26 RSV season | 4,407 (24.8%) | 665 (27.8%) | 3,742 (24.4%) |  |
| **Month tested** |  |  |  | 0.86 |
| Oct-Nov | 3,057 (17.2%) | 485 (20.3%) | 2,572 (16.7%) |  |
| Dec-Jan | 5,914 (33.3%) | 1,393 (58.3%) | 4,521 (29.4%) |  |
| Feb-Mar | 4,013 (22.6%) | 420 (17.6%) | 3,593 (23.4%) |  |
| Apr-Sep | 4,771 (26.9%) | 90 (3.8%) | 4,681 (30.5%) |  |
| **Gestational age (weeks)** |  |  |  | 0.12 |
| Median (IQR) | 39.0 (37.0, 39.0) | 39.0 (37.0, 39.0) | 39.0 (37.0, 39.0) |  |
| Missing | 4,487 (25.3%) | 674 (28.2%) | 3,813 (24.8%) |  |
| **Birth weight (g)** |  |  |  | 0.14 |
| Median (IQR) | 3,218.9 (2,819.0, 3,556.7) | 3,268.7 (2,909.1, 3,578.8) | 3,208.9 (2,809.1, 3,548.7) |  |
| Missing | 4,494 (25.3%) | 678 (28.4%) | 3,816 (24.8%) |  |
| **Insurance type** |  |  |  | 0.08 |
| Private | 5,835 (32.9%) | 854 (35.8%) | 4,981 (32.4%) |  |
| Public | 11,794 (66.4%) | 1,513 (63.4%) | 10,281 (66.9%) |  |
| Unknown | 126 (0.7%) | 21 (0.9%) | 105 (0.7%) |  |
| **Hospitalized<sup>4</sup>** | 2,841 (16.0%) | 495 (20.7%) | 2,346 (15.3%) | 0.14 |
| **Admitted to PICU<sup>4</sup>** | 584 (3.3%) | 49 (2.1%) | 535 (3.5%) | -0.09 |
| **Preterm** | 2,351 (17.6%) | 255 (14.7%) | 2,096 (18.0%) | -0.09 |
| Missing | 4,368 (24.6%) | 654 (27.4%) | 3,714 (24.2%) |  |
| **Cardiac diseases, including congenital heart diseases** | 1,163 (6.6%) | 96 (4.0%) | 1,067 (6.9%) | -0.13 |
| **Having at least one risk factor<sup>5</sup>** | 4,671 (26.3%) | 549 (23.0%) | 4,122 (26.8%) | -0.09 |
| **Ever immunized with nirsevimab** | 4,266 (24.0%) | 374 (15.7%) | 3,892 (25.3%) | -0.24 |
| **Immunized with nirsevimab during the surveillance year of RSV test** | 2,229 (12.6%) | 130 (5.4%) | 2,099 (13.7%) | -0.28 |
| **Mother received Abrisvo<sup>6</sup>** |  |  |  |  |
| Yes | 976 (11.1%) | 108 (10.8%) | 868 (11.1%) | -0.01 |
| No | 7,855 (44.2%) | 889 (37.2%) | 6,966 (45.3%) |  |
| Unknown<sup>6</sup> | 8,924 (50.3%) | 1,391 (58.2%) | 7,533 (49.0%) |  |

<sup>1</sup>n (%)
<sup>2</sup>Standardized Mean Difference. Covariates with an absolute standardized mean difference greater than 0.2 were considered to have important imbalances.
<sup>3</sup>Including American Indian or Native American, Asian, Middle Eastern or Northern African, and Pacific Islander by self-reporting.
<sup>4</sup>Admission within 14 days of the RSV test.
<sup>5</sup>Have at least one of the following conditions recorded in the infant's medical history or diagnosis records: (1) asthma, (2) immunodeficiency (e.g., transplantation history or leukemia), (3) cardiac diseases (including congenital heart diseases diagnosed at birth or any reporting of heart conditions), (4) pulmonary diseases, (5) Down syndrome, (6) small for gestational age (birth weight <2500 g), and (7) prematurity (gestational age <37 weeks).
<sup>6</sup>Unknown immunization status of the maternal RSV vaccine due to delivery of the child outside the YNHHS, and the immunization records of mothers couldn't be retrieved.

### RSV Activity and Nirsevimab Immunization by Season

RSV peaked at different times during the three RSV seasons. From 2023/24 to 2025/26, there was a progressively delayed peak and more gradual increase in the RSV positivity rate among the study population (Figure 1A). The positivity rate during 2025/26 did not begin increasing until mid-November and remained high compared with previous seasons through late February. In contrast to the delayed RSV activity, nirsevimab immunization started earlier in the latter two seasons and was predominantly administered during October to December, whereas immunization in the 2023/24 season continued throughout the entire RSV season (Figure 1B, 1C).

**Figure 1.**
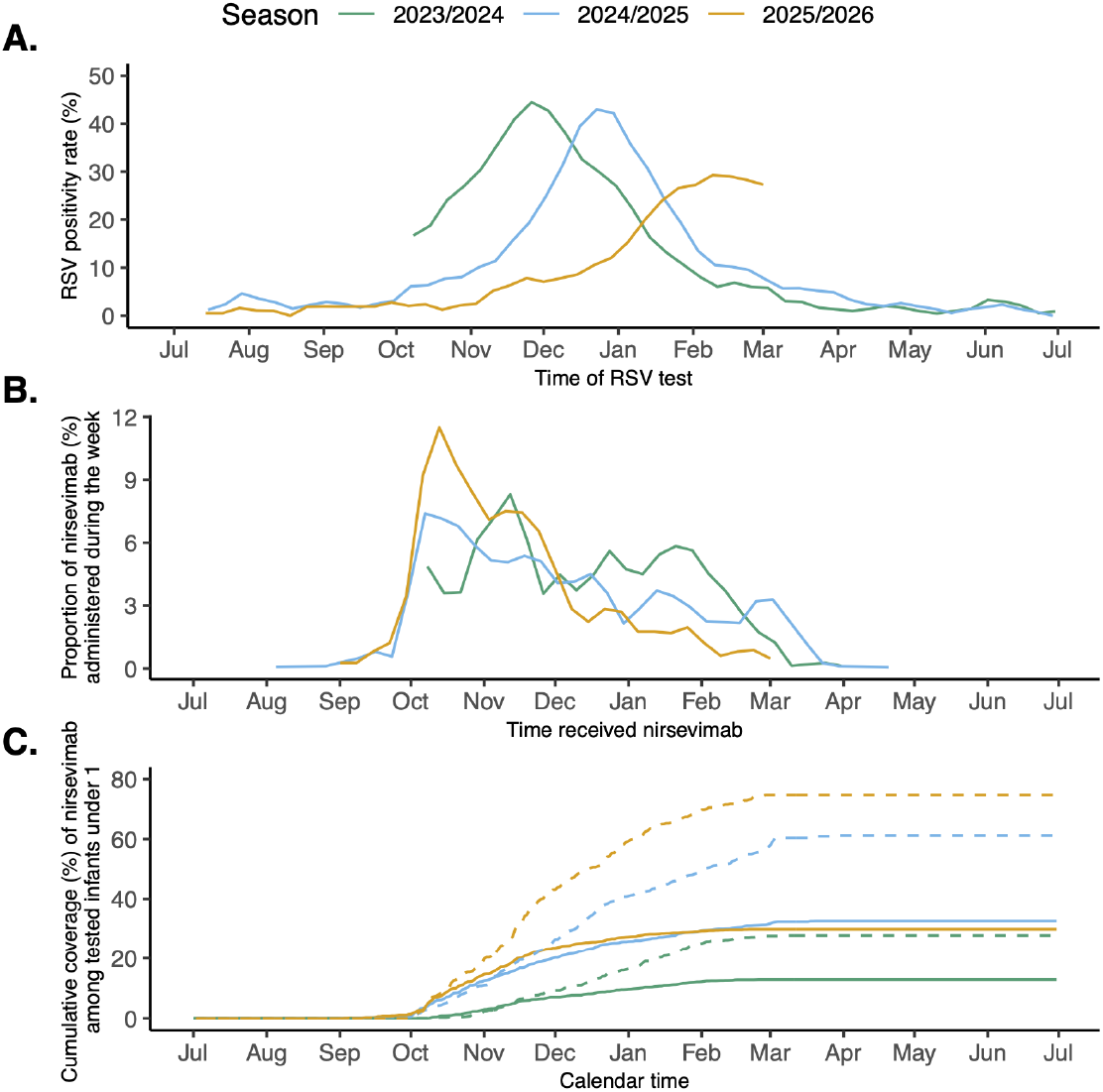
RSV activity and nirsevimab administration by surveillance year, October 1, 2023, to March 1, 2026. **A)** RSV positivity rate among tested patients over time for the three years. **B)** Timing of nirsevimab received during the three RSV seasons. The y-axis represents the proportion of nirsevimab doses administered in a specific week, calculated using the number of nirsevimab doses administered during a specific week divided by the total number of nirsevimab doses administered during that surveillance year (July-June). **C)** Cumulative coverage of nirsevimab. Solid line: cumulative coverage among all infants under 1 year of age at time of RSV test who were eligible for nirsevimab during the RSV season (i.e., born during RSV season or aged under 8 months at the start of October), capturing the coverage of both birth dose and catch-up dose. Dashed lines: cumulative coverage among only infants under 1 year of age born during the RSV season (October-March), capturing only the coverage of birth dose.

### Effectiveness of Nirsevimab by Surveillance Year

In the primary analysis, we included 9,681 tested children, of whom 3,859, 4,026, and 1,796 were from the 2023/24, 2024/25, and 2025/26, respectively. Comparing the analytic population by surveillance year, the median age at the time of the test for RSV was 6.6, 6.1, and 4.6 months for 2023/24, 2024/25, and 2025/26, and the median time interval from nirsevimab immunization to a positive RSV test increased from 43 days in 2023/24, to 65 days in 2024/25, and to 87 days in 2025/26 (Table S3). Pooling data from the three years, the overall effectiveness of nirsevimab was 67.3% (95% confidence interval [CI]: 59.8, 73.3%) against medically attended RSV infection, 60.2% (95% CI: 49.6, 68.5%) against RSV-associated outpatient visits and 88.9% (95% CI: 82.3, 93.0%) against RSV-associated hospitalization (Figure 2).

**Figure 2.**
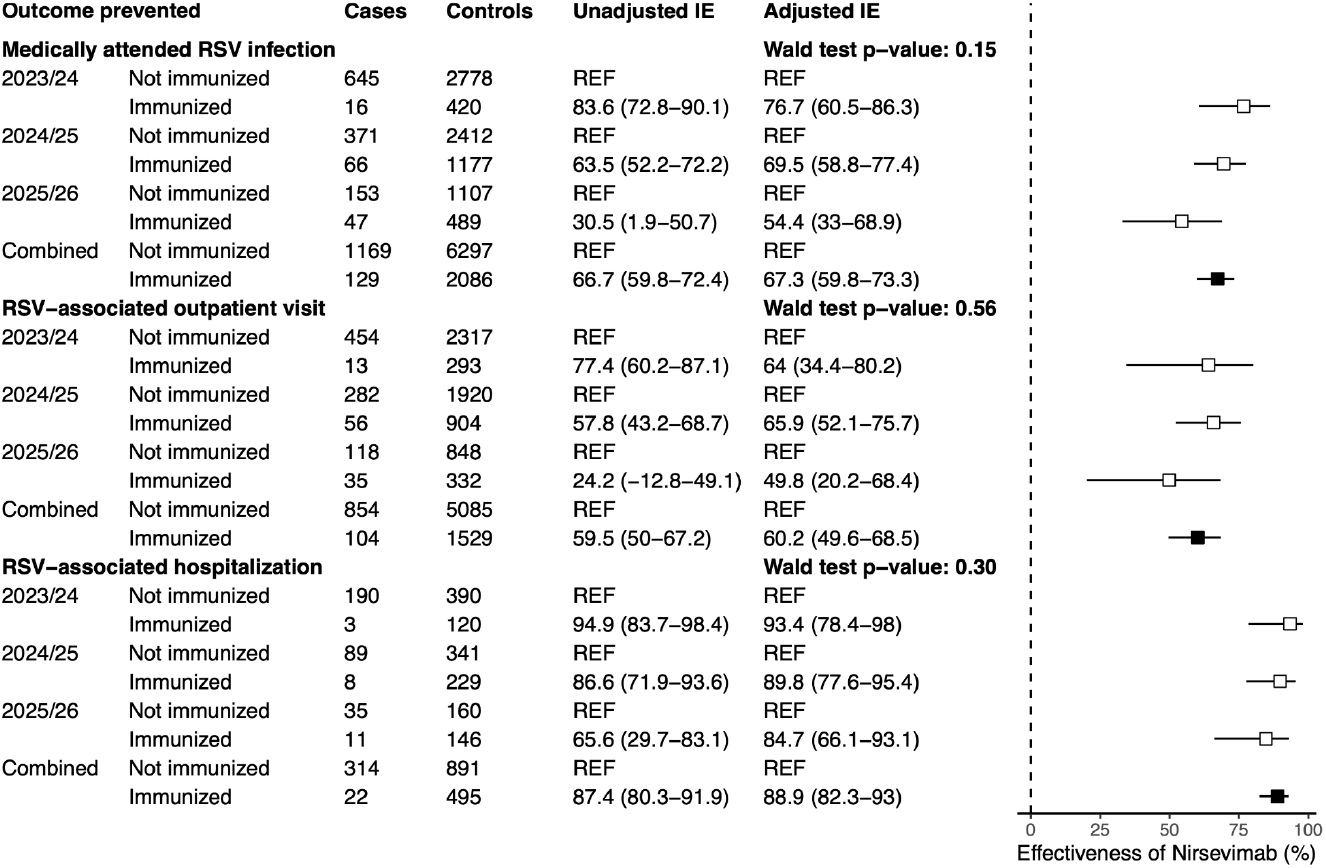
Estimated overall and stratified effectiveness of nirsevimab against medically attended RSV infection by encounter setting and surveillance year. The square points and error bars indicate the medians and 95% confidence intervals (CI) of the estimates. Effectiveness across years were compared and tested using Wald test and p-values are shown for each disease outcome. Adjusted model controlled for age at RSV test, RSV activity (log-transformed weekly RSV positivity rate in Connecticut, obtained from the National Syndromic Surveillance Program (NSSP) via PopHIVE [16]), presence of at least one risk factor for severe RSV disease, and maternal RSV vaccine status.

Across the three examined clinical outcomes, point estimates of the overall effectiveness were lower in 2025/26 compared with 2023/24 and 2024/25 (Figure 2). The overall effectiveness against medically attended RSV infection was estimated to be 76.7%, 69.5%, and 54.4% during 2023/24, 2024/25, and 2025/26, respectively. The overall effectiveness against RSV-associated hospitalization also declined across the three years, but with a more modest decrease (93.4%, 89.8%, and 84.7%).

Importantly, early effectiveness within the first 2 months post immunization did not differ significantly across surveillance years (Wald test p-value: 0.63), and the pattern of declining point estimates observed for infection and outpatient visits was no longer observed when restricting to this early period, with estimates of 81.8%, 73.5%, and 76.5% against medically attended RSV infection in the first 2 months post immunization in 2023/24, 2024/25, and 2025/26, respectively (Figure S3). Sensitivity analyses generated consistent trends in both overall and initial effectiveness across years (Figure S4, S5).

### Waning Protection from Nirsevimab

The estimated effectiveness of nirsevimab waned over the first 12 months after immunization and plateaued thereafter (Figure 3). Pooling data from three seasons, effectiveness against RSV infection declined from 79.3% (95% CrI: 70.8, 85.9%) at 1 month post immunization to 65.3% (95% CrI: 57.3, 72.0%) at 3 months, to 42.7% (95% CrI: 25.5, 55.6%) at 6 months, to 26.4% (95% CrI: 2.6, 42.4%) at 9 months, and to 18.6% (95% CrI: -0.5, 33.5%) at 12 months post immunization. Estimated effectiveness against RSV-associated hospitalization declined more slowly, from 92.5% (95% CrI: 85.9, 96.4%) at 1 month, to 88.1% (95% CrI: 80.7, 93.1%) at 3 months, to 77.2% (95% CrI: 60.4, 87.6%) at 6 months, to 60.2% (95% CrI: 31.4, 77.4%) at 9 months, and to 39.9% (95% CrI: 2.4, 63.3%) at 12 months post immunization. Waning curves were similar for 2024/25 and 2025/26, with similar shapes and starting values (Figure 3, S7). Estimates for 2023/24 showed much wider uncertainty intervals; this corresponded to the smaller number of patients with a long interval between nirsevimab immunization and RSV test in this year compared with the other two years (Figure S8). Estimates of waning effectiveness from the alternative model structures generated consistent declining patterns (Figure S9-11).

**Figure 3.**
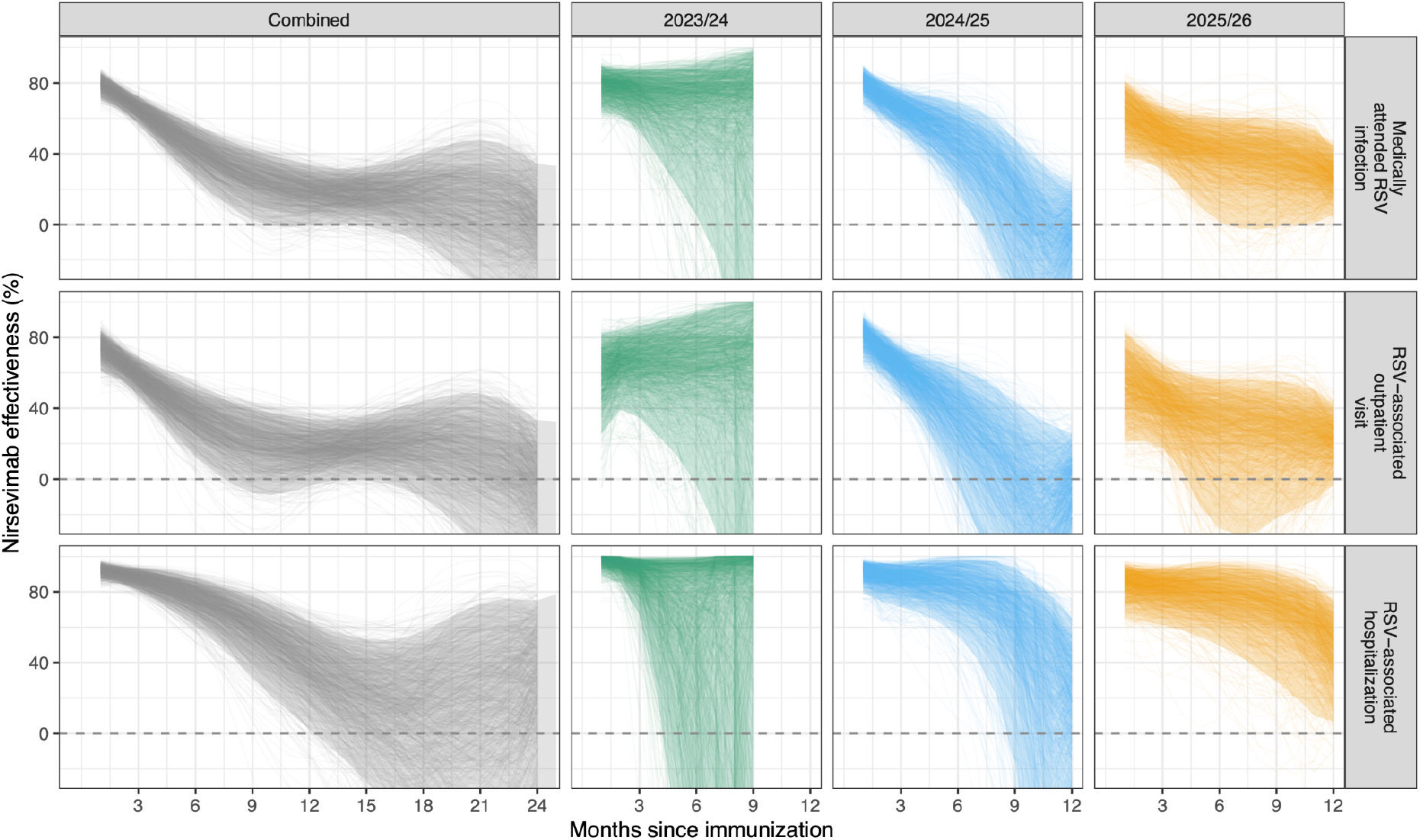
Estimated effectiveness of nirsevimab over time since immunization in preventing medically attended RSV infection, by encounter setting and surveillance year. We randomly sampled 1,000 iterations from the posterior of each model; each line represents one sampled trajectory. The shaded areas represent 95% credible intervals. For the unstratified analysis (combining data from all three years), we analysed nirsevimab effectiveness up to 24 months post immunization. Analysis was conducted up to 12 months when stratified by surveillance year as sample sizes are small for time-since-immunization intervals beyond 12 months. Among infants tested during 2023/24 (the second column), no infants were immunized more than 9 months before the RSV test. Models were adjusted for age at RSV test, RSV activity (log-transformed weekly RSV positivity rate in Connecticut, obtained from the National Syndromic Surveillance Program (NSSP) via PopHIVE [16]), presence of at least one risk factor for severe RSV diseases, and maternal RSV vaccine status.

## Discussion

In this test-negative case-control study using EHR data collected from three consecutive seasons, we found that nirsevimab was effective in protecting infants and young children against medically attended RSV-associated infections. Pooling data from three seasons, our estimates of effectiveness were similar to those of other post-licensure studies [8,9,11,18]. We estimated a lower point estimate for the effectiveness in 2025/26, which corresponded to successively delayed RSV activity from 2023/24 to 2025/26. Our analysis showed that protection from nirsevimab waned over time during the first 12 months, and the rate of waning was slower against hospitalization, with effectiveness sustained at 60–80% through 6 to 9 months post immunization. Nirsevimab’s effectiveness beyond 12 months after immunization plateaued and protection was limited, with high uncertainty in the estimates.

Our study makes several contributions to the existing literature on the effectiveness of nirsevimab. To our knowledge, this is the first study characterizing the temporal dynamic of nirsevimab’s effectiveness over an extended follow-up period including more than 12 months after immunization. With the data collection period spanning three seasons, we were able to capture more patients with longer intervals after immunization with nirsevimab, and we assessed the effectiveness beyond 6 months after immunization with more precision than in our previous study [10]. Our finding that protection from nirsevimab against RSV-associated hospitalization remained high (60-80%) at 6 to 9 months after immunization, aligned with another recent study from the US, which reported an effectiveness of 77% at 130 to 210 days post immunization [11]. Results from our analysis also provided insight into the protection from nirsevimab during the infants’ second RSV season. Evidence on the effectiveness of immunization during the second season has been sparse and inconsistent across studies. A recent US study reported no significant protection, with a point estimate of 8% against RSV-associated hospitalization in the second season [11], while a study from Spain reported a statistically significant 55% effectiveness against RSV-associated LRTI hospitalization in the second season [19]. Estimates from our analysis, including monthly estimates of effectiveness beyond 12 months post immunization, indicate that a low effectiveness of nirsevimab might be sustained into the second season, but there is still considerable uncertainty in the estimates. The plateau in effectiveness between 12 months and 18 months might indicate some degree of immune boosting among immunized infants exposed to RSV during their second season. However, this positive plateau with high uncertainty beyond 12 months should be interpreted cautiously, as it could also reflect residual confounding rather than true biological protection.

To our knowledge, this is the first study to evaluate how timing of RSV epidemics could influence the effectiveness of nirsevimab. We report and compare individual-level effectiveness across three consecutive years. With the approval of nirsevimab in 2023, this study provides timely real-world data on its effectiveness over multiple years following its introduction. There have been concerns that specific amino acid substitutions in the pre-F fusion protein could lead to reduced effectiveness of nirsevimab. While genomic surveillance suggests these mutations are rare, the increasing coverage of nirsevimab over time could lead to increased selective pressures on the viral population [20]. Alternatively, variation in RSV epidemic timing could affect the degree of waning immunity between immunization and peak RSV exposure. We found that in Connecticut, nirsevimab’s effectiveness was lower for all examined outcomes in 2025/26, compared with the previous two years. This coincided with delayed RSV activity and an increasing median time interval from nirsevimab immunization to a positive RSV test over the three years. Therefore, protection from nirsevimab had, on average, waned more in 2025/26 by the time RSV activity peaked. However, our models with interaction terms between surveillance year and time since immunization showed that initial effectiveness in the first 2 months after immunization was comparable across the three years. Meanwhile, waning effectiveness for the latter seasons had similar shapes and starting values. These indicate that the lower effectiveness in 2025/26 was more likely due to the longer duration between immunization and exposure to RSV rather than an overall reduction in effectiveness due to emergence of escape variants. The smaller variation in effectiveness against hospitalization across years was also consistent with the estimated slower rate of waning in effectiveness against hospitalization. Our findings suggest that effectiveness of nirsevimab depends on the timing of administration relative to the timing of the RSV season. This underlines the importance of aligning the nirsevimab administration schedule with local RSV activity, as more flexible and targeted timing relative to the predicted RSV peaks could enhance its overall protection.

Our study has several limitations. First, under-ascertainment of risk factors and symptoms might occur due to incomplete documentation within the EHR system. To address this, we applied a comprehensive set of free-text searches and ICD-10-CM codes to maximize the capture of the documented information. Second, in our analyses of the waning effectiveness of nirsevimab, unmeasured confounders such as undocumented RSV infections between immunization and the RSV test might have occurred. However, this is unlikely to substantially bias our estimates of nirsevimab’s effectiveness against RSV-associated hospitalization, since infants with more severe respiratory symptoms are unlikely to remain undetected. Third, as nirsevimab likely confers “leaky” (i.e., partial) immunity when the protection wanes, the differential depletion of susceptible individuals in the immunized and unimmunized groups may lead to underestimation of effectiveness and overestimation of the rate of waning immunity [21]. Fourth, RSV vaccination status of mothers may have been incompletely ascertained for infants without linked maternal records in our study population. However, our sensitivity analysis imputing the missing vaccine records of mothers generated estimates similar to our main results.

## Conclusion

In this test-negative case-control study across three consecutive RSV seasons, nirsevimab provided sustained protection against RSV-associated hospitalization through 6 to 12 months after immunization, but protection beyond 12 months was limited. The lower point estimates of nirsevimab’s effectiveness in the 2025/26 was consistent for both hospitalized and non-hospitalized patients with RSV infections and coincided with delayed RSV activity, highlighting the importance of aligning nirsevimab administration with local RSV circulation.

## Data Availability

All data produced in the present study are available upon reasonable request to the authors. External researchers can make written requests to the corresponding author for sharing of completely de-identified and aggregate-level data.

https://github.com/Hanmeng-Xu/season2_RSV_risk

