## Supplementary Materials for "Durability and Seasonal Variation in the Effectiveness of Nirsevimab over Three Seasons in Connecticut"

#### Supplementary methods

##### Estimating the effectiveness of nirsevimab by time since immunization

We evaluated the waning protection from nirsevimab by time since immunization using a logistic regression model within a Bayesian framework. For the $j$th test record in our dataset, the observed case status (i.e., whether the patient tested positive or negative for RSV) followed a Bernoulli distribution, such that

$$Case\_Status_j \sim Bernoulli(p_j).$$

The time between nirsevimab immunization and RSV testing among those immunized individuals was categorized into bi-weekly intervals. To incorporate this variable in the regression model, we created indicator variables to represent each time category ( $time\_since\_vax_{jn}$, $n = 1, 2, 3...$). For example, if an individual $j$ was immunized 0-2 weeks before testing, then $time\_since\_vax_{j1} = 1$ and $time\_since\_vax_{jn} = 0$ for $n = 2, 3, ....$ For an unimmunized individual, all the indicator variables $time\_since\_vax_{jn}$ took the value of 0. The probability of an individual testing positive for RSV $p_j$, was modeled using a multivariable logistic regression framework as follows:

$$logit(p_j) = \beta_0 + \sum_{n=1}^{N} \beta_n time\_since\_vax_{jn} + \sum_{m=1}^{M} \alpha_m Z_{jm}$$

where $\beta_n$ is the immunization effect coefficient for each time since immunization category ( $time\_since\_vax_n$, $n = 1, 2, 3...N$, in which $N$ represents the total number of time since immunization categories), and $Z_{jm}$ represents potential confounders, in which $m$ represents the number of confounders included in the regression model. We included age at testing, time tested (RSV season vs. off-season), RSV activity (RSV positivity rate), presence of at least one risk factor for severe RSV disease, and whether the mother was immunized with the maternal RSV vaccine as confounders in the model, and $\alpha_m$'s represent the coefficients for the confounders.

We tested waning models with different hierarchical structures for the effectiveness coefficients $\beta_n$, including the model using basis spline (B-spline) structure in our main analysis, and models with an imposed monotonic trend or autoregressive structures in our sensitivity analyses, described below.

**Model 1: B-Spline model:**

In the B-spline model, the effectiveness coefficient $\beta_n$ was modeled as a linear combination of different "trends" over time since immunization:

$$\beta_n = \gamma_1 B_{n,1} + \gamma_2 B_{n,2} + .... + \gamma_k B_{n,k}$$

in which $B$ is the basis spline matrix with a degree of freedom of $k$ and $\gamma$'s are the spline coefficients.  This ensured smoothness in the effectiveness estimates. An example of $B$ matrix with a degree of freedom of 6 is shown below:

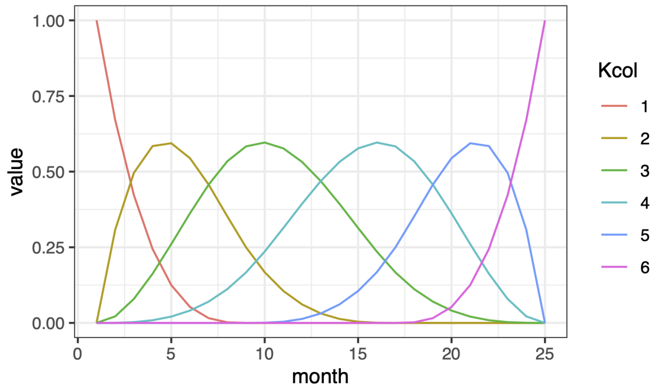


Each colored line ($B_k$) represents a trend with a specific shape over time and the estimated $\beta_n$ over time is a linear combination of these trends. We adopted a random walk structure on the spline coefficients $\gamma$'s:

$$\gamma_1 \sim N(0,\ 100^2)$$

$$\gamma_k \sim N(\gamma_{k-1},\ \tau_{spl}^{-1})$$

$$\tau_{spl} \sim Gamma(0.01,\ 0.01)$$

We tested models with the degree of freedom taking the values of 4,5, and 6, and compared the Deviance Information Criterion (DIC) across models. The difference in DIC across models was less than 2 ($DIC_{k=4}$=1949, $DIC_{k=5}$=1949, $DIC_{k=6}$=1948, against RSV-associated hospitalization, pooling records from the three years). We used the model with the degree of freedom of 6 in our main analysis.

**Model 2: Imposed a monotonic trend on $\beta_n$:**

Given the waning nature of passive immunity, we assume that nirsevimab effectiveness follows a non-increasing trend over time post immunization. This structure allowed relatively lower flexibility compared to the other models, allowing $\beta_n$ to either increase or remain unchanged over time since immunization, such that effectiveness could only decrease or plateau over time post immunization. To reflect this in the model, we imposed a monotonic structure on the regression coefficients $\beta_n$'s, such that

$$\beta_{n+1} = \beta_n + d_{n+1},\ n = 1, 2, 3, \ldots$$

$$d_{n+1} \sim N(0,\ \tau_d^{-1})T(0,\ \infty)$$

$$\tau_d \sim Gamma(0.01,\ 0.01)$$

$T(0,\ \infty)$ represents truncation at 0, allowing $d_{n+1}$ to take only non-negative values, and we used weakly informative priors on the increment $d_n$ over time. For the first coefficient $\beta_1$ (coefficient for the effectiveness 0-2 weeks after immunization), we also used a weakly informative prior distribution:

$$\beta_1 \sim N(0,\ 100^2)$$

**Model 3: Autoregressive $\beta_n$:**

We also adopted a model structure without any imposed trend in the effectiveness over time since immunization, allowing the model to estimate the effectiveness coefficients with more flexibility. Instead, we added a first-order autoregressive structure (i.e., AR(1)) on the coefficients to prevent abrupt changes between neighboring coefficients:

$$\beta_1 \sim N(0,\ [\tau_{ar}(1 - \rho_{ar}^2)]^{-1})$$

$$\beta_n \sim N(\rho_{ar}\beta_{n-1},\ \tau_{ar}^{-1}),\ n = 2, 3, 4....$$

$$\rho_{ar} \sim Uniform(-1,\ 1)$$

$$\tau_{ar} \sim Gamma(0.01,\ 0.01)$$

**Model 4: Autoregressive $\beta_n$ with linear mean function:**

On the basis of model 3 with only the AR(1) structure in $\beta_n$, we extended the AR(1) prior on the immunization effect over time by incorporating a linear mean function, such that:

$$\beta_1 \sim N(0,\ [\tau_{arL}(1 - \rho_{arL}^2)]^{-1})\ \text{(same as Model 3)}$$

$$\beta_n \sim N(\mu_0 + \mu_1(n - 1) + \rho_{arL}\beta_{n-1},\ \tau_{arL}^{-1}),\ n = 2, 3, 4....$$

$$\mu_0, \mu_1 \sim N(0,\ 100^2)$$

$$\rho_{arL} \sim Uniform(-1,\ 1)$$

$$\tau_{arL} \sim Gamma(0.01,\ 0.01)$$

This structure passed a weakly informative trends to the $\beta_n$ over time, and could stabilize the estimates over time more compared to Model 3 without the linear mean structure.

We evaluated the waning effectiveness of nirsevimab using the above models and examined the effectiveness over time against various clinical endpoints, including medically attended RSV infection, RSV-associated outpatient visits, and RSV-associated hospitalization. Models for each endpoint were fitted separately via the rjags package in R version 4.3.1, in which we collected 10,000 samples from the posterior distribution after discarding the first 50,000 samples in the burn-in period. Convergence was evaluated using trace plots (Figure S6). The estimated effectiveness of nirsevimab after a given period of time (for the time interval $n$) since immunization $IE_n$ was calculated as

$$IE_n = \left(1 - e^{\beta_n}\right) * 100\%, \ n = 1, 2, 3, ...$$

Medians and 95% quantile-based credible intervals were calculated from the collected posterior samples.

### Supplementary Figures

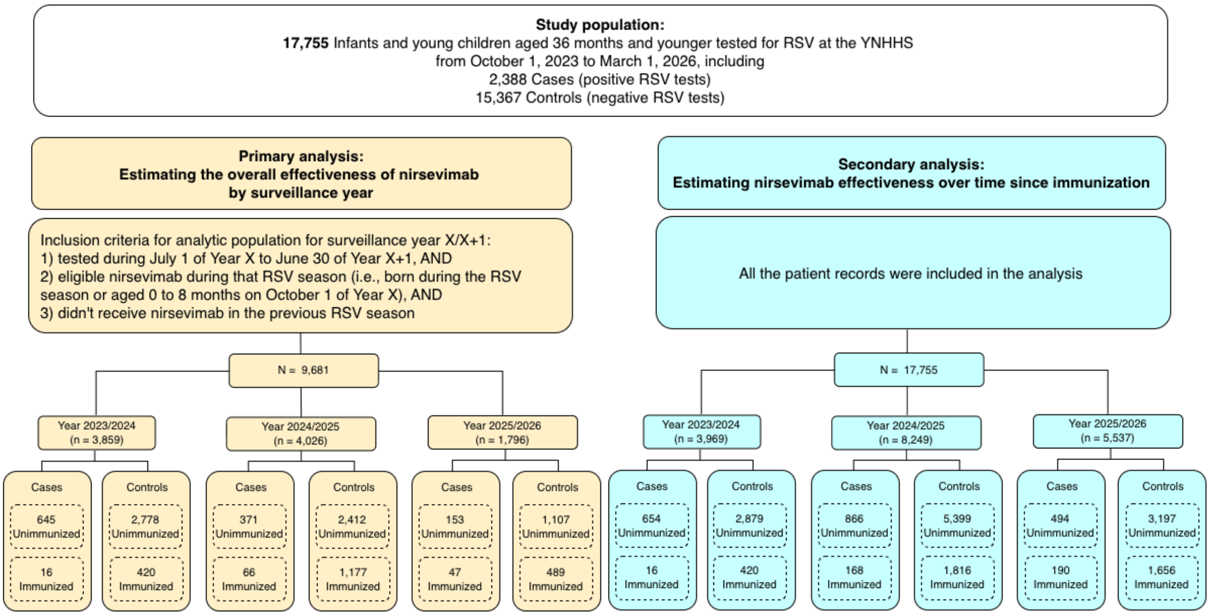


**Figure S1. Overview of the eligibility criteria and sample size for the analyses.**

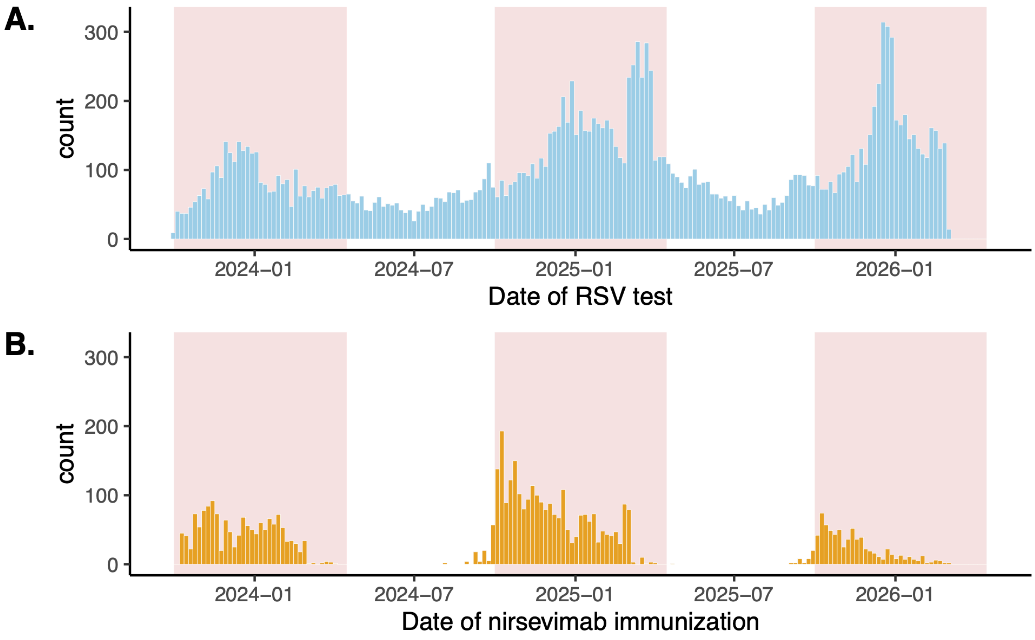


**Figure S2. A) Overview of the included RSV tests during the study period, and B) Timing of nirsevimab administration among the tested records.** The rectangles in salmon color indicate the three consecutive RSV seasons (October-March, from 2023/24 to 2025/26) covered by the study period. RSV testing was conducted year-round, both within and outside RSV season.

| Outcome prevented | | Cases | Controls | Unadjusted IE | Adjusted IE |
|---|---|---|---|---|---|
| **Medically attended RSV infection** | | | | | **Wald test p−value: 0.63** |
| 2023/24 | Not immunized | 645 | 2778 | REF | REF |
| | Immunized <= 2m | 10 | 170 | 74.7 (51.8–87) | 81.8 (64.8–90.6) |
| 2024/25 | Not immunized | 371 | 2412 | REF | REF |
| | Immunized <= 2m | 30 | 418 | 53.3 (31.3–68) | 73.5 (60.1–82.4) |
| 2025/26 | Not immunized | 153 | 1107 | REF | REF |
| | Immunized <= 2m | 14 | 292 | 65.3 (39.1–80) | 75.3 (56–86.1) |
| Combined | Not immunized | 1169 | 6297 | REF | REF |
| | Immunized <= 2m | 54 | 880 | 66.9 (56.6–75.3) | 76.5 (68.6–82.8) |
| **RSV−associated outpatient visit** | | | | | **Wald test p−value: 0.73** |
| 2023/24 | Not immunized | 454 | 2317 | REF | REF |
| | Immunized <= 2m | 9 | 103 | 55.4 (11.2–78) | 69.7 (38.3–85.2) |
| 2024/25 | Not immunized | 282 | 1920 | REF | REF |
| | Immunized <= 2m | 22 | 267 | 43.9 (11.8–64) | 71.7 (54.2–82.5) |
| 2025/26 | Not immunized | 118 | 848 | REF | REF |
| | Immunized <= 2m | 7 | 180 | 72.1 (39.1–87) | 79.7 (55–90.8) |
| Combined | Not immunized | 854 | 5085 | REF | REF |
| | Immunized <= 2m | 38 | 550 | 58.9 (43.2–71.1) | 72.9 (61.5–81.4) |
| **RSV−associated hospitalization** | | | | | **Wald test p−value: 0.25** |
| 2023/24 | Not immunized | 190 | 390 | REF | REF |
| | Immunized <= 2m | 1 | 61 | 96.6 (75.6–100) | 97.2 (79.3–99.6) |
| 2024/25 | Not immunized | 89 | 341 | REF | REF |
| | Immunized <= 2m | 6 | 121 | 81 (55.4–92) | 88.9 (72.9–95.4) |
| 2025/26 | Not immunized | 35 | 160 | REF | REF |
| | Immunized <= 2m | 6 | 102 | 73.1 (33.8–89) | 82.1 (54.5–93) |
| Combined | Not immunized | 314 | 891 | REF | REF |
| | Immunized <= 2m | 13 | 284 | 87 (77.9–93) | 90.4 (83.3–95) |

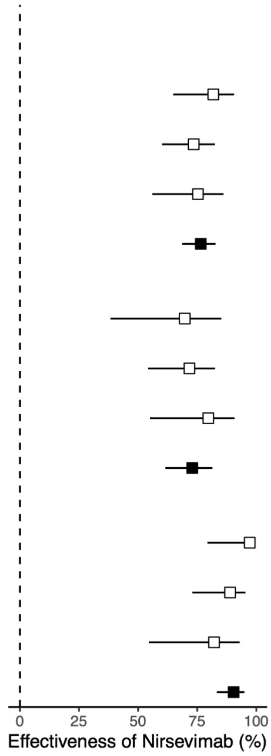


**Figure S3. Effectiveness of nirsevimab in the first 2 months after immunization, by disease severity and by surveillance year.** In this analysis, we only included records immunized within 2 months before the RSV test as "immunized". The square points and error bars indicate the medians and 95% confidence intervals (CI) of the estimates. We compared the effectiveness across years: The null hypothesis that effectiveness was homogeneous across all seasons was evaluated using a joint Wald test, in which the interaction terms between exposure and surveillance year were tested simultaneously equal to zero (implemented via the *linearHypothesis* function in the *car* package in R). A p-value < 0.05 was considered evidence of statistically significant heterogeneity in effectiveness across years. Adjusted model controlled for age at RSV test, RSV activity (log-transformed weekly RSV positivity rate in Connecticut, obtained from the National Syndromic Surveillance Program (NSSP) via PopHIVE [19]), presence of at least one risk factor for severe RSV disease, and maternal RSV vaccine status.

| Sensitivity analysis | | Cases | Controls | Unadjusted IE | Adjusted IE |
|---|---|---|---|---|---|
| **SA1: exclude those eligible for nirsevimab in the previous season** | | | | | **Wald test p–value: 0.16** |
| 2023/24 | Not immunized | 645 | 2778 | REF | REF |
| | Immunized | 16 | 420 | 83.6 (73.7–90.5) | 76.7 (61.7–86.8) |
| 2024/25 | Not immunized | 282 | 1896 | REF | REF |
| | Immunized | 63 | 1122 | 62.2 (50.2–71.8) | 69.4 (58.4–77.8) |
| 2025/26 | Not immunized | 126 | 922 | REF | REF |
| | Immunized | 46 | 473 | 28.8 (−0.8–50.6) | 54.6 (33.3–69.5) |
| **SA2: exclude patients coinfected w/ RSV and other resp. viruses** | | | | | **Wald test p–value: 0.16** |
| 2023/24 | Not immunized | 643 | 2778 | REF | REF |
| | Immunized | 16 | 420 | 83.5 (73.6–90.5) | 76.7 (61.6–86.8) |
| 2024/25 | Not immunized | 356 | 2412 | REF | REF |
| | Immunized | 63 | 1177 | 63.7 (52.5–72.7) | 69.9 (59.3–78) |
| 2025/26 | Not immunized | 146 | 1107 | REF | REF |
| | Immunized | 45 | 489 | 30.2 (1.7–51.3) | 54.5 (33.2–69.5) |
| **SA3: restrict controls to those positive for other resp. viruses** | | | | | **Wald test p–value: 0.20** |
| 2023/24 | Not immunized | 645 | 203 | REF | REF |
| | Immunized | 16 | 34 | 85.2 (73–92.2) | 84 (66.5–92.6) |
| 2024/25 | Not immunized | 371 | 457 | REF | REF |
| | Immunized | 66 | 248 | 67.2 (55.8–76) | 71.8 (60.1–80.3) |
| 2025/26 | Not immunized | 153 | 272 | REF | REF |
| | Immunized | 47 | 137 | 39 (10.8–58.8) | 58.2 (35.7–73.2) |
| **SA4: exclude whose vaccinated with MV or with unknown MV status** | | | | | **Wald test p–value: 0.19** |
| 2023/24 | Not immunized | 114 | 604 | REF | REF |
| | Immunized | 9 | 191 | 75 (52.5–88.4) | 64.5 (29.7–83.9) |
| 2024/25 | Not immunized | 225 | 1498 | REF | REF |
| | Immunized | 46 | 921 | 66.7 (54.3–76.3) | 74.8 (64.4–82.5) |
| 2025/26 | Not immunized | 94 | 742 | REF | REF |
| | Immunized | 37 | 375 | 22.1 (−15.2–48.3) | 53.8 (29–70.4) |
| **SA5: adjust for calendar month instead of RSV positivity rate** | | | | | **Wald test p–value: 0.02** |
| 2023/24 | Not immunized | 645 | 2778 | REF | REF |
| | Immunized | 16 | 420 | 83.6 (73.7–90.5) | 74.1 (57.2–85.3) |
| 2024/25 | Not immunized | 371 | 2412 | REF | REF |
| | Immunized | 66 | 1177 | 63.5 (52.5–72.4) | 65.2 (53–74.5) |
| 2025/26 | Not immunized | 153 | 1107 | REF | REF |
| | Immunized | 47 | 489 | 30.5 (2.7–51.1) | 54.2 (33–69.1) |
| **SA6: impute missing MV status** | | | | | **Wald test p–value: 0.02** |
| 2023/24 | Not immunized | 645 | 2778 | REF | REF |
| | Immunized | 16 | 420 | 83.6 (73.7–90.5) | 74.7 (58.2–85.7) |
| 2024/25 | Not immunized | 371 | 2412 | REF | REF |
| | Immunized | 66 | 1177 | 63.5 (52.5–72.4) | 65.6 (53.8–74.7) |
| 2025/26 | Not immunized | 153 | 1107 | REF | REF |
| | Immunized | 47 | 489 | 30.5 (2.7–51.1) | 54.6 (33.9–69.3) |

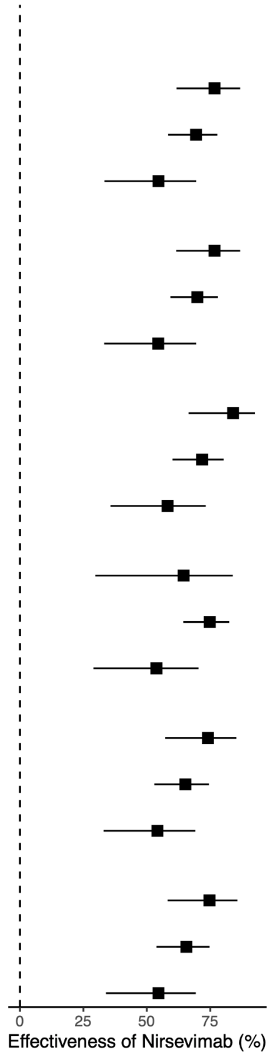


**Figure S4. Sensitivity analysis (SA) of the overall effectiveness of nirsevimab, by disease severity and by surveillance year.** The square points and error bars represent the median and 95% confidence intervals of the adjusted estimates. In SA1, infants eligible for nirsevimab in the previous RSV season included those born during the previous season or aged under 8 months at the start of October of the previous RSV season. In SA2 and SA3, "other respiratory viruses" included influenza, SARS-CoV-2, and human metapneumovirus (hMPV). In SA5, values were imputed for participants with unknown maternal RSV vaccination status (~50%, due to delivery outside of YNHHS) by random sampling from the three other categories (vaccinated within 6 months, vaccinated over 6 months, unvaccinated) using the proportions observed among those with known maternal RSV vaccine status.

| Sensitivity analysis | | Cases | Controls | Unadjusted IE | Adjusted IE |
|---|---|---|---|---|---|
| **SA1: exclude those eligible for nirsevimab in the previous season** | | | | | **Wald test p–value: 0.58** |
| 2023/24 | Not immunized | 645 | 2778 | REF | REF |
| | Immunized <= 2m | 10 | 170 | 74.7 (51.8–87) | 82 (65.1–90.7) |
| 2024/25 | Not immunized | 282 | 1896 | REF | REF |
| | Immunized <= 2m | 29 | 403 | 51.6 (28–67) | 72.8 (58.8–82.1) |
| 2025/26 | Not immunized | 126 | 922 | REF | REF |
| | Immunized <= 2m | 14 | 284 | 63.9 (36.3–80) | 74.9 (55.1–85.9) |
| **SA2: exclude patients coinfected w/ RSV and other resp. viruses** | | | | | **Wald test p–value: 0.57** |
| 2023/24 | Not immunized | 643 | 2778 | REF | REF |
| | Immunized <= 2m | 10 | 170 | 74.6 (51.6–87) | 81.9 (65–90.6) |
| 2024/25 | Not immunized | 356 | 2412 | REF | REF |
| | Immunized <= 2m | 30 | 418 | 51.4 (28.4–67) | 72.6 (58.7–81.8) |
| 2025/26 | Not immunized | 146 | 1107 | REF | REF |
| | Immunized <= 2m | 14 | 292 | 63.6 (36.1–79) | 74.3 (54.2–85.5) |
| **SA3: restrict controls to those positive for other resp. viruses** | | | | | **Wald test p–value: 0.37** |
| 2023/24 | Not immunized | 645 | 203 | REF | REF |
| | Immunized <= 2m | 10 | 10 | 68.5 (23.3–87) | 85.5 (61.7–94.5) |
| 2024/25 | Not immunized | 371 | 457 | REF | REF |
| | Immunized <= 2m | 30 | 68 | 45.7 (14.7–65) | 69.5 (49.6–81.5) |
| 2025/26 | Not immunized | 153 | 272 | REF | REF |
| | Immunized <= 2m | 14 | 68 | 63.4 (32.7–80) | 77.1 (56.8–87.9) |
| **SA4: exclude whose vaccinated with MV or with unknown MV status** | | | | | **Wald test p–value: 0.96** |
| 2023/24 | Not immunized | 114 | 604 | REF | REF |
| | Immunized <= 2m | 6 | 86 | 63 (13.4–84) | 72.9 (35–88.7) |
| 2024/25 | Not immunized | 225 | 1498 | REF | REF |
| | Immunized <= 2m | 23 | 313 | 51.1 (23.6–69) | 74.2 (58.3–84) |
| 2025/26 | Not immunized | 94 | 742 | REF | REF |
| | Immunized <= 2m | 10 | 220 | 64.1 (30–82) | 76.7 (53.4–88.3) |
| **SA5: adjust for calendar month instead of RSV positivity rate** | | | | | **Wald test p–value: 0.49** |
| 2023/24 | Not immunized | 645 | 2778 | REF | REF |
| | Immunized <= 2m | 10 | 170 | 74.7 (51.8–87) | 80.2 (61.9–89.7) |
| 2024/25 | Not immunized | 371 | 2412 | REF | REF |
| | Immunized <= 2m | 30 | 418 | 53.3 (31.3–68) | 69.2 (53.9–79.4) |
| 2025/26 | Not immunized | 153 | 1107 | REF | REF |
| | Immunized <= 2m | 14 | 292 | 65.3 (39.1–80) | 75.8 (57.2–86.3) |
| **SA6: impute missing MV status** | | | | | **Wald test p–value: 0.50** |
| 2023/24 | Not immunized | 645 | 2778 | REF | REF |
| | Immunized <= 2m | 10 | 170 | 74.7 (51.8–87) | 80.8 (63.1–90) |
| 2024/25 | Not immunized | 371 | 2412 | REF | REF |
| | Immunized <= 2m | 30 | 418 | 53.3 (31.3–68) | 70.2 (55.5–80.1) |
| 2025/26 | Not immunized | 153 | 1107 | REF | REF |
| | Immunized <= 2m | 14 | 292 | 65.3 (39.1–80) | 76.2 (57.9–86.5) |

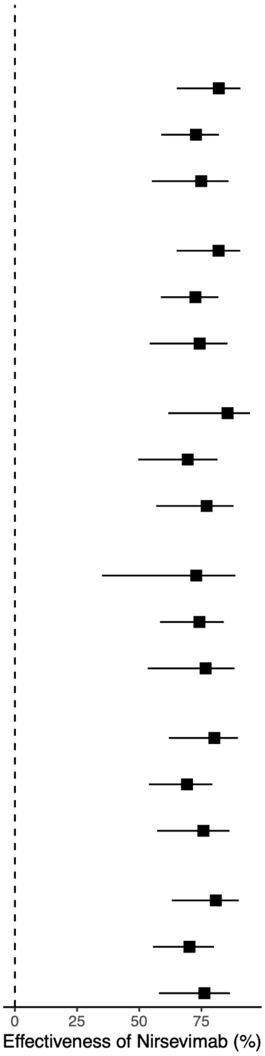


**Figure S5. Sensitivity analysis (SA) of the nirsevimab effectiveness in the first 2 months after immunization, by disease severity and by surveillance year.** In this analysis, we only included records immunized within 2 months before the RSV test as "immunized". The square points and error bars represent the median and 95% confidence intervals of the adjusted estimates. In SA1, infants eligible for nirsevimab in the previous RSV season included those born during the previous season or aged under 8 months at the start of October of the previous RSV season. In SA2 and SA3, "other respiratory viruses" included influenza, SARS-CoV-2, and human metapneumovirus (hMPV). In SA5, values were imputed for participants with unknown maternal RSV vaccination status (~50%, due to delivery outside of YNHHS) by random sampling from the three other categories (vaccinated within 6 months, vaccinated over 6 months, unvaccinated) using the proportions observed among those with known maternal RSV vaccine status.

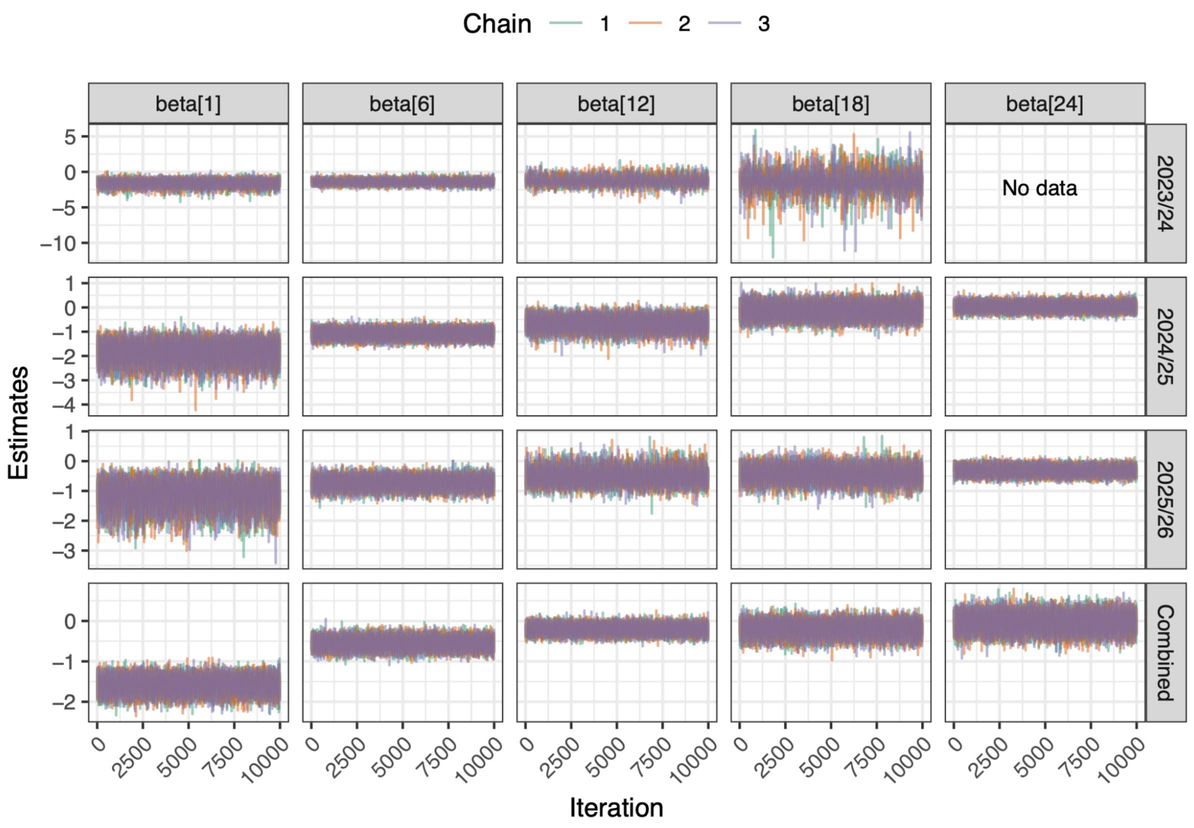


**Figure S6. Trace plots for the coefficients of the waning effectiveness of nirsevimab over time since immunization.** Trace plots of effectiveness coefficients derived from the spline model, both by surveillance year and pooling three years' data. For the by-year analysis (the first three rows), we presented the trace plots of five out of twenty-five effectiveness coefficients (betas), which represent the coefficients at 2 weeks, 3, 6, 9, and 12 months post the immunization of nirsevimab. For the analysis pooling the three surveillance years (the last row), the analysis was conducted up to 24 months and we also presented the trace plots of five out of twenty-five effectiveness coefficients (betas), which represent the coefficients at 1, 6, 12, 18, and 24 months post immunization. Iterations of the burn-in period (50,000 burn-in iterations) were discarded and not plotted here, and only the 10,000 sampled iterations are plotted.

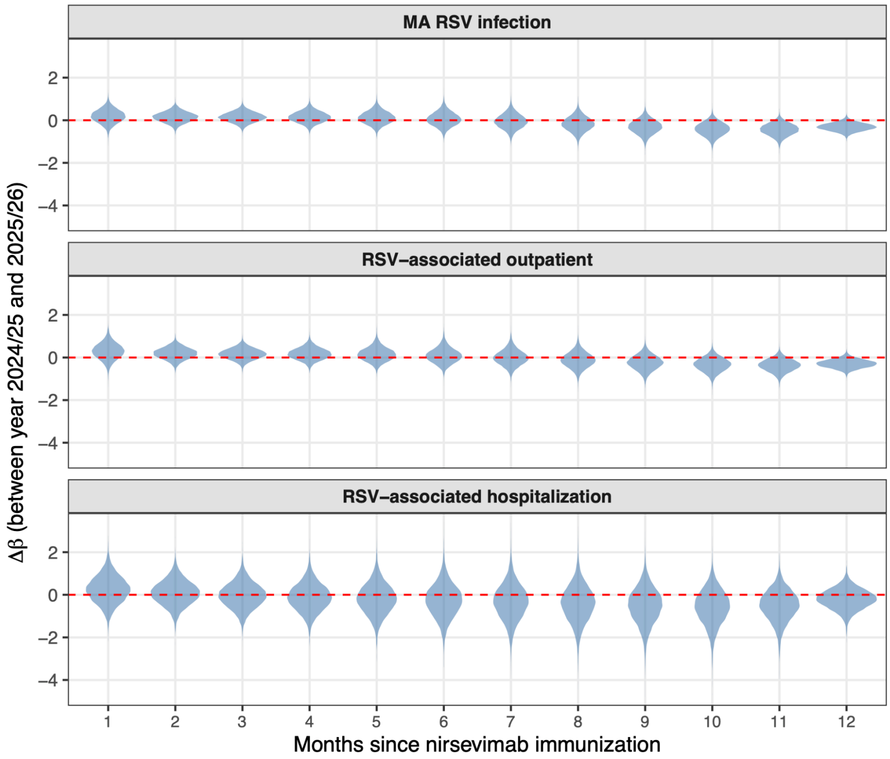


**Figure S7. Posterior distributions of the difference in nirsevimab effectiveness (β coefficients) between surveillance year 2024/25 and 2025/26, by time since immunization and clinical setting.** Here, we estimated the effectiveness over time pooling records from all three years in one single model with the spline structure, and Δβ was calculated as $\beta_{2025/26} - \beta_{2024/25}$ for each time-since-immunization interval. Each violin represents the posterior distribution of Δβ. The red dashed line indicates no between-season difference (Δβ = 0). All the Δβ's have 95% credible intervals (CrI) crossing 0, indicating no significant difference in the effectiveness over time between 2024/25 and 2025/26. We calculated the difference between 2025/26 and 2024/25, but didn't use estimates from the 2023/24 season, as sample size for 2023/24 was much smaller (Figure 8) and estimates had much wider uncertainty levels (Figure 3).

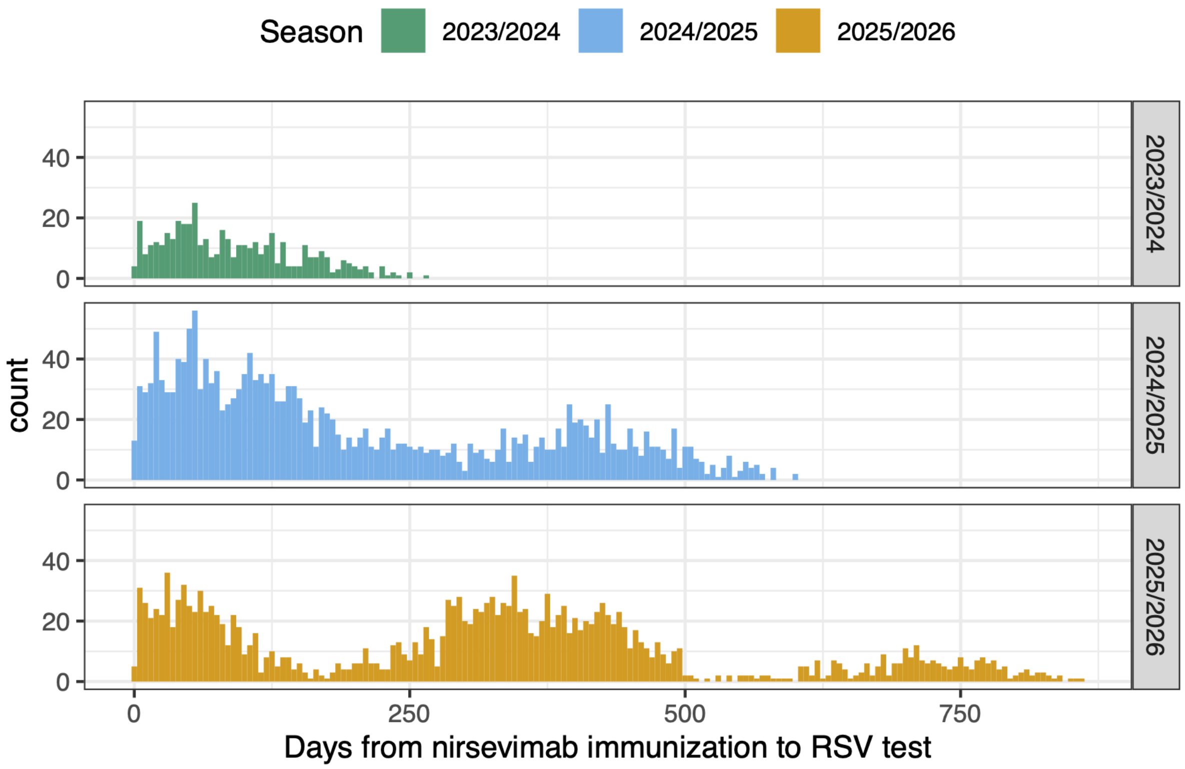


**Figure S8. Distribution of time elapsed from nirsevimab immunization to the RSV test among children immunized with nirsevimab included in the model estimating the waning effectiveness.** The RSV season during surveillance year 2023/24 was the first RSV season of nirsevimab administration, and there were substantially fewer patients with longer time intervals due to fewer patients being tested towards the end of the RSV season, compared with the subsequent two years.

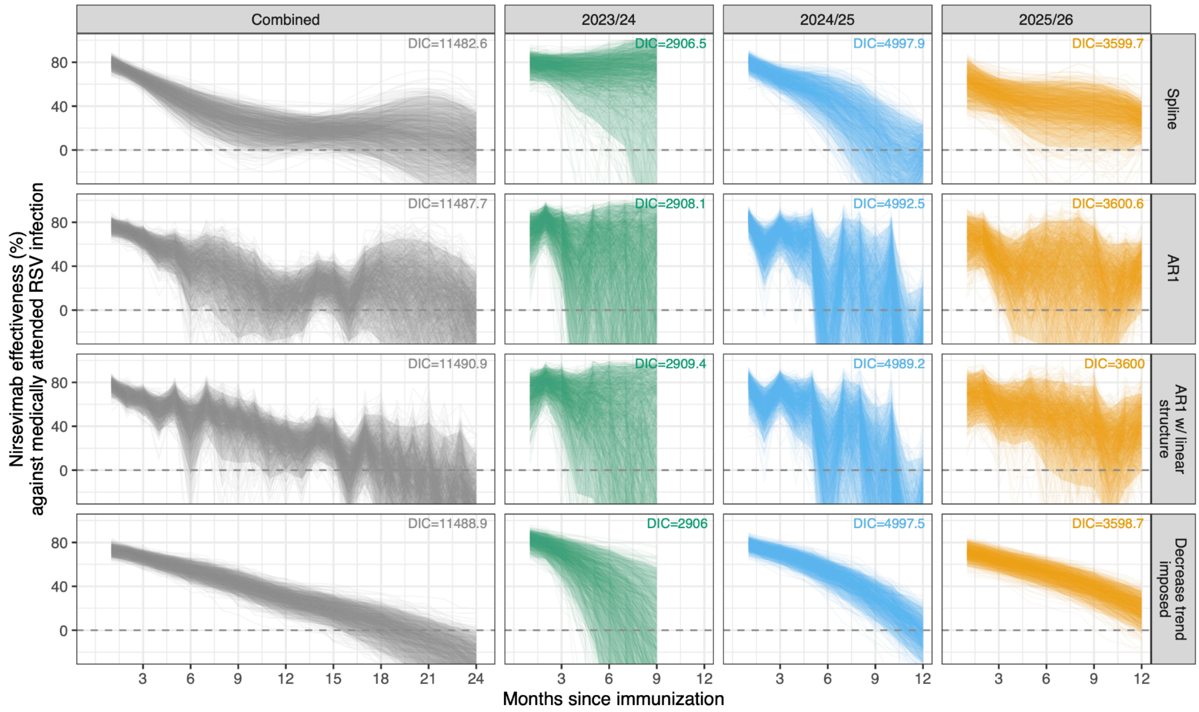


**Figure S9. Nirsevimab effectiveness against medically attended RSV infection over time since immunization, estimated from models with alternative structures**. We reported results from the Spline model (the first row) as our results in the main text (Figure 3), and here we compare the main results with estimates from models with alternative structures. We randomly sampled 1,000 iterations from the posterior of each model; each line represents one sampled trajectory. The shaded areas represent 95% credible intervals. We present the waning curves both stratified by surveillance year (2023/24, 2024/25, 2025/26) and pooling data from three years ("Combined"). Analysis was conducted up to 12 months for the stratified analysis as sample sizes are small for time-since-immunization intervals beyond 12 months. The unstratified analysis was run up to 24 months post immunization. DIC value (Deviance Information Criterion) for each model was shown in each panel. A full description of all the model structures is in the Supplementary methods section above. Among infants tested during the 2023/24 surveillance year (the second column), the uncertainty is large due to the small sample size, and no infants were immunized more than 9 months before the RSV test.

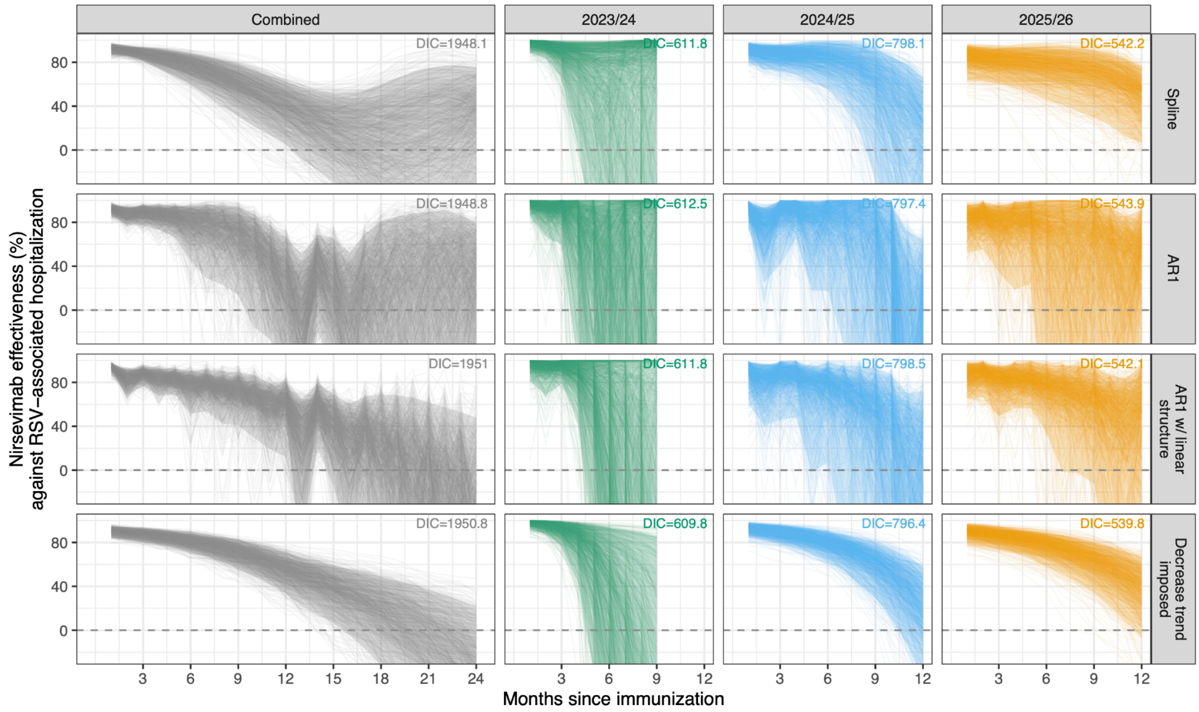


**Figure S10. Nirsevimab effectiveness against RSV-associated hospitalization over time since immunization, estimated from models with alternative structures**. We reported results from the Spline model (the first row) as our results in the main text (Figure 3), and here we compare the main results with estimates from models with alternative structures. We randomly sampled 1,000 iterations from the posterior of each model; each line represents one sampled trajectory. The shaded areas represent 95% credible intervals. We present the waning curves both stratified by surveillance year (2023/24, 2024/25, 2025/26) and pooling data from three years ("Combined"). Analysis was conducted up to 12 months for the stratified analysis as sample sizes are small for time-since-immunization intervals beyond 12 months. The unstratified analysis was run up to 24 months post immunization. DIC value (Deviance Information Criterion) for each model was shown in each panel. A full description of all the model structures is in the Supplementary methods section above. Among infants tested during the 2023/24 surveillance year (the second column), the uncertainty is large due to the small sample size, and no infants were immunized more than 9 months before the RSV test.

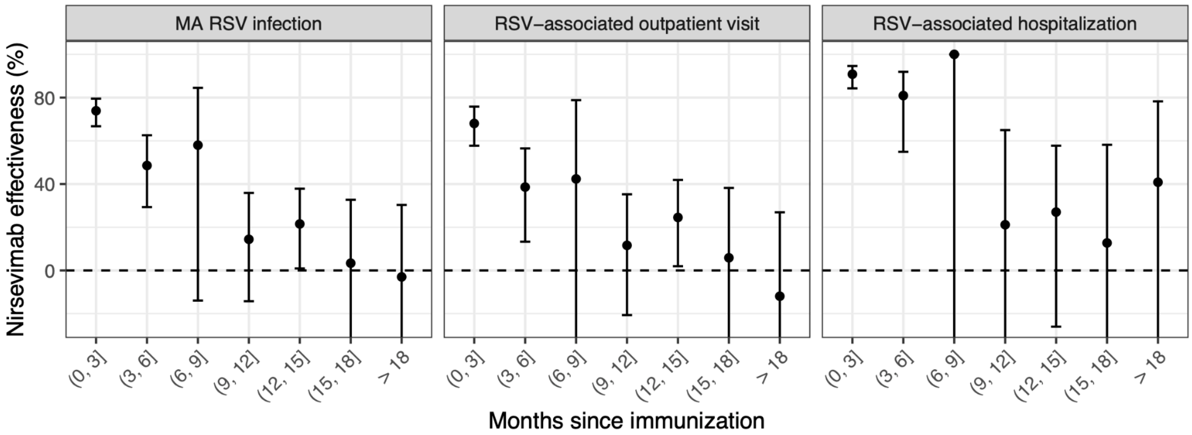


**Figure S11. Nirsevimab effectiveness estimated from a multivariable logistic regression model, stratifying by the time from immunization to the RSV test.** The points and error bars represent medians and 95% confidence intervals of the estimates. The figure shows adjusted effectiveness, controlled for the same set of confounders as in the Bayesian models, including age at test, RSV positivity during the week of the RSV test (log-transformed weekly RSV positivity rate in Connecticut, obtained from the National Syndromic Surveillance Program (NSSP) via PopHIVE [1]), presence of at least one risk factor for severe RSV disease, and immunization status of the maternal RSV vaccine.

#### Supplementary Tables

##### Table S1. Free-text patterns and ICD-10-CM codes used to identify risk factors for severe RSV diseases and respiratory symptoms.

| Category | Target item | Identification codes and free-text patterns |
|---|---|---|
| Risk factor for severe RSV disease | Prematurity | Premature, prematurity, extreme immaturity of newborn, extremely low birth weight, preterm, preterm newborn, preterm birth, 15-36 completed weeks of gestation, 15-36 weeks gestation, P07.3, P07.2<br>*(In addition to medical history and delivery records, we also used gestational age to classify prematurity, defined as gestational age < 37 weeks.)* |
| | Low birth weight | Low birth weight, extremely low birth weight, less than 500 grams, 500-749 grams, 750-999 grams, 1,000-1,249 grams, 1,250-1,499 grams, 1,500-1,749 grams, 1,750-1,999 grams, 2,000-2,249 grams, 2,250-2,499 grams, P07.0, P07.1<br>*(In addition to medical history and delivery records, we also used birth weight to classify low birth weight, defined as birth weight < 2,500 grams.)* |
| | Asthma | Asthma, reactive airway disease, J45 |
| | Congenital heart disease | Congenital heart disease, coarctation of aorta, congenital heart, D-TGA, double outlet right ventricle, dextrocardia, DORV, cardiac abnormality, ventricular septa, mitral regurgitation, coarctation of aorta, hypoplastic aortic arch, atrial septal defect, heart abnormality, cardiomegaly, patent ductus arteriosus, tetralogy of fallot, atrioventricular canal (AVC), persistent left superior vena cava, aortic arch anomaly, wolff-Parkinson-White (WPW) syndrome, supraventricular tachycardia, partial AV canal, atrial septa, supraventricular tachycardia, partial anomalous pulmonary venous return, aortic coarctation, congenital coronary artery fistula to pulmonary artery, peripheral pulmonic stenosis, tricuspid atresia with normal great arteries, Q20, Q21, Q22, Q23, Q24, Q25 |
| | Immunodeficiency | Liver transplanted, transplant, leukemia, D80, D81, D82, D83, D84, Z94, B20, B21, B22, B23, B24, C91, C92, C93, C94, C95 |
| | Down syndrome | Down syndrome, trisomy 21, Q90 |
| | Chronic lung disease of prematurity | Bronchopulmonary dysplasia, P27.1, P27.0, P27.8, P27.9 |
| | Having at least one risk factor | *Defined as having at least one of the above risk factors.* |
| Respiratory symptoms | Fever | Fever, febrile, R50 |
| | Cough | Cough, R05 |
| | Wheezing | Wheezing, R06.2 |
| | Difficulty breathing | Abnormal breathing, breathing difficulty, difficulty breathing, shortness of breath, breathing problem, dyspnea, apnea, tachypnea, R06 |
| | Upper respiratory tract infection (URTI) | Abnormal breathing, breathing difficulty, difficulty breathing, breathing problem, shortness of breath, cough, acute obstructive laryngitis, pharyngitis, otitis media, upper respiratory infection, upper respiratory tract infection, upper resp. tract infection, upper resp. infection, croup, nasopharyngitis, pain in throat, reactive airway disease, nasal congestion, sore throat, URI, laryngeal stridor, J00, J01, J02, J03, J04, J05, J06 |
| | Lower respiratory tract infection (LRTI) | Bronchiolitis, bronchitis, laryngotracheobronchitis, bronchospasm, acute chest syndrome, acute hypoxic respiratory failure, pneumonia, hypoxia, hypoxemia, lower resp. tract infection, lower resp. infection, lower respiratory tract infection, lower respiratory infection, hypoxemic respiratory failure, acute respiratory failure with hypoxia, wheezing, J12, J13, J14, J15, J16, J17, J18, J20, J21, J22 |

**Table S2. Comparison of included patients by nirsevimab immunization status prior to the RSV test.** "Immunized" was defined as receiving nirsevimab before the RSV test, regardless of season of immunization.

| Characteristic | **Overall**, N = 17,755[1] | **Immunized**, N = 4,266[1] | **Unimmunized**, N = 13,489[1] | SMD[2] |
|---|---|---|---|---|
| **Sex** | | | | 0.01 |
| Female | 7,685 (43.3%) | 1,859 (43.6%) | 5,826 (43.2%) | |
| Male | 10,067 (56.7%) | 2,407 (56.4%) | 7,660 (56.8%) | |
| Missing | 3 (0.02%) | 0 | 3 (0.02%) | |
| **Race and ethnicity** | | | | 0.22 |
| Black non-Hispanic | 3,303 (18.6%) | 917 (21.5%) | 2,386 (17.7%) | |
| White non-Hispanic | 4,360 (24.6%) | 887 (20.8%) | 3,473 (25.7%) | |
| Other non-Hispanic[3] | 1,067 (6.0%) | 249 (5.8%) | 818 (6.1%) | |
| Hispanic | 8,043 (45.3%) | 2,090 (49.0%) | 5,953 (44.1%) | |
| unknown | 982 (5.5%) | 123 (2.9%) | 859 (6.4%) | |
| **Age at RSV test (months)** | | | | -0.32 |
| Mean (SD) | 11.5 (8.0) | 9.7 (6.6) | 12.1 (8.3) | |
| Median (IQR) | 10.1 (5.3, 16.5) | 9.0 (4.2, 13.7) | 10.4 (5.7, 17.6) | |
| **Age at nirsevimab immunization (month)** | | | | |
| Mean (SD) | 1.9 (2.5) | 1.9 (2.5) | NA | NA |
| Median (IQR) | 0.4 (0.0, 3.5) | 0.4 (0.0, 3.5) | NA | |
| **Time of test** | | | | 0.54 |
| 2023/24 RSV season | 3,391 (19.1%) | 356 (8.3%) | 3,035 (22.5%) | |
| 2024 off-season | 1,713 (9.6%) | 197 (4.6%) | 1,516 (11.2%) | |
| 2024/25 RSV season | 6,212 (35.0%) | 1,622 (38.0%) | 4,590 (34.0%) | |
| 2025 off-season | 2,032 (11.4%) | 552 (12.9%) | 1,480 (11.0%) | |
| 2025/26 RSV season | 4,407 (24.8%) | 1,539 (36.1%) | 2,868 (21.3%) | |
| **Month tested** | | | | 0.24 |
| Oct-Nov | 3,057 (17.2%) | 543 (12.7%) | 2,514 (18.6%) | |
| Dec-Jan | 5,914 (33.3%) | 1,579 (37.0%) | 4,335 (32.1%) | |
| Feb-Mar | 4,013 (22.6%) | 1,169 (27.4%) | 2,844 (21.1%) | |
| Apr-Sep | 4,771 (26.9%) | 975 (22.9%) | 3,796 (28.1%) | |
| **Gestational age (weeks)** | | | | -0.25 |
| Median (IQR) | 39.0 (37.0, 39.0) | 38.0 (37.0, 39.0) | 39.0 (37.0, 39.0) | |
| Missing | 4,487 (25.3%) | 381 (8.9%) | 4,106 (30.4%) | |
| **Birth weight (g)** | | | | -0.22 |
| Median (IQR) | 3,218.9 (2,819.0, 3,556.7) | 3,129.0 (2,630.4, 3,508.8) | 3,248.9 (2,879.1, 3,568.9) | |
| Missing | 4,494 (25.3%) | 388 (9.1%) | 4,106 (30.4%) | |
| **Insurance type** | | | | 0.26 |
| Private | 5,835 (32.9%) | 1,049 (24.6%) | 4,786 (35.5%) | |
| Public | 11,794 (66.4%) | 3,208 (75.2%) | 8,586 (63.7%) | |
| Unknown | 126 (0.7%) | 9 (0.2%) | 117 (0.9%) | |
| **Hospitalized[4]** | 2,841 (16.0%) | 851 (19.9%) | 1,990 (14.8%) | 0.14 |
| **Admitted to PICU[4]** | 584 (3.3%) | 187 (4.4%) | 397 (2.9%) | 0.08 |
| **Preterm** | 2,351 (17.6%) | 954 (24.4%) | 1,397 (14.7%) | 0.25 |
| Missing | 4,368 | 356 | 4,012 | |
| **Cardiac diseases, including congenital heart diseases** | 1,163 (6.6%) | 418 (9.8%) | 745 (5.5%) | 0.16 |
| **Having at least one risk factor[5]** | 4,671 (26.3%) | 1,496 (35.1%) | 3,175 (23.5%) | 0.26 |
| **Mom received abrysvo** | 976 (11.1%) | 559 (16.4%) | 417 (7.7%) | 0.27 |
| Unknown[6] | 8,924 (50.3%) | 854 (20.0%) | 8,070 (59.8%) | |
| **Difficulty breathing** | 1,157 (6.5%) | 425 (10.0%) | 732 (5.4%) | 0.17 |
| **Cough** | 2,208 (12.4%) | 493 (11.6%) | 1,715 (12.7%) | -0.04 |
| **Fever** | 4,980 (28.0%) | 1,187 (27.8%) | 3,793 (28.1%) | -0.01 |
| **Symptoms in LRT** | 3,995 (22.5%) | 946 (22.2%) | 3,049 (22.6%) | -0.01 |
| **Wheezing** | 359 (2.0%) | 90 (2.1%) | 269 (2.0%) | 0.01 |

[1]n (%)
[2]Standardized Mean Difference. Covariates with an absolute standardized mean difference greater than 0.2 were considered to have important imbalances.
[3]Including American Indian or Native American, Asian, Middle Eastern or Northern African, and Pacific Islander by self-reporting.
[4]Admitted to hospital or PICU within 14 days of the RSV test.
[5]Have at least 1 of the following conditions recorded in the infant's medical history or diagnosis records: (1) asthma, (2) immunodeficiency (e.g., transplantation history or leukemia), (3) cardiac diseases (including congenital heart diseases diagnosed at birth or any reporting of

heart conditions), (4) pulmonary diseases, (5) Down syndrome, (6) small for gestational age (birth weight <2500 g), and (7) prematurity (gestational age <37 weeks).
[6] Unknown immunization status of the maternal vaccine due to delivery of the child outside the YNHHS, and the immunization records of mothers couldn't be retrieved.

#### Table S3. Comparison of characteristics of the analytic population by RSV season included for the nirsevimab effectiveness analysis.

| Characteristic | **Overall**, N = 9,681[1] | **Season 2023/24**, N = 3,859[1] | **Season 2024/25**, N = 4,026[1] | **Season 2025/26**, N = 1,796[1] |
|---|---|---|---|---|
| **RSV test results** | | | | |
| Cases | 1,298 (13.4%) | 661 (17.1%) | 437 (10.9%) | 200 (11.1%) |
| Controls | 8,383 (86.6%) | 3,198 (82.9%) | 3,589 (89.1%) | 1,596 (88.9%) |
| **Sex (female)** | 4,188 (43.3%) | 1,658 (43.0%) | 1,719 (42.7%) | 811 (45.2%) |
| **Race and ethnicity** | | | | |
| Black non-Hispanic | 1,701 (17.6%) | 707 (18.3%) | 702 (17.4%) | 292 (16.3%) |
| White non-Hispanic | 2,516 (26.0%) | 1,044 (27.1%) | 1,011 (25.1%) | 461 (25.7%) |
| Other non-Hispanic[2] | 569 (5.9%) | 231 (6.0%) | 231 (5.7%) | 107 (6.0%) |
| Hispanic | 4,328 (44.7%) | 1,684 (43.6%) | 1,828 (45.4%) | 816 (45.4%) |
| unknown | 567 (5.9%) | 193 (5.0%) | 254 (6.3%) | 120 (6.7%) |
| **Age at RSV test (months, median, IQR)** | 5.9 (3.0, 8.8) | 6.6 (3.5, 9.4) | 6.1 (3.2, 9.0) | 4.6 (2.0, 7.1) |
| **Gestation (weeks, median, IQR)** | 39.0 (37.0, 39.0) | 39.0 (37.0, 39.0) | 39.0 (37.0, 39.0) | 39.0 (37.0, 39.0) |
| Missing | 2,151 (22.2%) | 846 (21.9%) | 903 (22.4%) | 402 (22.4%) |
| **Birth weight (g, median, IQR)** | 3,218 (2,799, 3,568) | 3,233.9 (2,849, 3,568) | 3,208 (2,749, 3,556) | 3,199.0 (2,799, 3,578) |
| Missing | 2,133 (22.0%) | 845 (21.9%) | 899 (22.3%) | 389 (21.7%) |
| **Insurance type** | | | | |
| private | 3,232 (33.4%) | 1,306 (33.8%) | 1,287 (32.0%) | 639 (35.6%) |
| public | 6,394 (66.0%) | 2,539 (65.8%) | 2,719 (67.5%) | 1,136 (63.3%) |
| unknown | 55 (0.6%) | 14 (0.4%) | 20 (0.5%) | 21 (1.2%) |
| **Hospitalized[3]** | 1,722 (17.8%) | 703 (18.2%) | 667 (16.6%) | 352 (19.6%) |
| **Admitted to PICU[3]** | 409 (4.2%) | 147 (3.8%) | 163 (4.0%) | 99 (5.5%) |
| **Preterm (gestation < 37 weeks)** | 1,360 (17.9%) | 529 (17.4%) | 597 (19.0%) | 234 (16.7%) |
| Missing | 2,089 (21.6%) | 818 (21.2%) | 879 (21.3%) | 392 (21.8%) |
| **Cardiac diseases, including congenital heart diseases** | 764 (7.9%) | 277 (7.2%) | 327 (8.1%) | 160 (8.9%) |
| **Having at least one risk factor[4]** | 2,482 (25.6%) | 1,047 (27.1%) | 1,033 (25.7%) | 402 (22.4%) |
| **Nirsevimab immunized[5]** | 2,215 (22.9%) | 436 (11.3%) | 1,243 (30.9%) | 536 (29.8%) |
| **Mother received Abrysvo[5]** | 498 (9.3%) | 100 (9.8%) | 328 (10.9%) | 70 (5.3%) |
| Unknown[6] | 4,327 (44.7%) | 2,841 (73.6%) | 1,008 (25.0%) | 478 (26.6%) |
| **Days from nirsevimab immunization to RSV test (Median, IQR)** | 69 (45, 101) | 43 (29, 90) | 65 (50, 100) | 87 (46, 103) |

[1]n (%)
[2]Including American Indian or Native American, Asian, Middle Eastern or Northern African, and Pacific Islander by self-reporting.
[3]Admitted to hospital or PICU within 14 days of the RSV test.
[4]Have at least 1 of the following conditions recorded in the infant's medical history or diagnosis records: (1) asthma, (2) immunodeficiency (e.g., transplantation history or leukemia), (3) cardiac diseases (including congenital heart diseases diagnosed at birth or any reporting of heart conditions), (4) pulmonary diseases, (5) Down syndrome, (6) small for gestational age (birth weight <2500 g), and (7) prematurity (gestational age <37 weeks).
[5]For the by-season overall effectiveness comparison, all the immunized individuals were immunized within the same surveillance year as the RSV test.
[6] Unknown immunization status of the maternal vaccine due to delivery of the child outside the YNHHS, and the immunization records of mothers couldn't be retrieved.
